# A survey-based study investigating opinions on genetic research among Swedish autistic individuals and parents of autistic children

**DOI:** 10.64898/2025.12.02.25341212

**Authors:** Samuelle Fajutrao Falk, Anna Hellquist, Kristiina Tammimies

**Affiliations:** Center of Neurodevelopmental Disorders, Centre for Psychiatry Research, Department of Women’s and Children’s Health, Karolinska Institutet, Solna, Sweden; Department of Highly Specialized Pediatric Orthopedics and Medicine, Astrid Lindgren Children’s Hospital, Karolinska University Hospital, Region Stockholm, Stockholm, Sweden; North Stockholm Psychiatry, Region Stockholm, Stockholm, Sweden, Department of Clinical Genetics, Akademiska Hospital, Region Uppsala, Uppsala, Sweden

**Keywords:** Autism, genetic research, attitudes, opinions, autistic individuals, parents of autistic individuals

## Abstract

The opinions of the autism community are crucial for the future of genetic research. This study examines the attitudes of autistic adolescents, adults, and parents of autistic individuals toward genetic research in Sweden. We aimed to determine respondents’ views on genetic research in general, and to understand their expectations and concerns. For this aim, we conducted two online surveys with closed-ended and open-ended questions, one aimed at parents of autistic children and another for autistic adolescents and adults. A total of 871 parents and 213 autistics participated. We show that the attitudes towards genetic research are generally positive, with both respondent groups hoping that it will lead to improved interventions, enhanced quality of life, better educational opportunities, and increased support services. The autistic group valued research studies that would provide individual results from genetic research. However, thematic analysis of the respondents’ concerns on genetic research revealed that there are significant concerns about the potential misuse of genetic information, particularly regarding eugenics. Our results underscore the importance of engaging the autism community in genetic research to ensure its relevance and ethical integrity, ultimately facilitating the translation of research outcomes into tangible benefits for individuals with autism and their families.

**Lay abstract:** Autistic adolescents, adults, and parents of autistic individuals generally have a positive attitude towards genetic research in autism. However, many are concerned about the potential misuse of the research results. We surveyed 871 parents of autistic individuals and 213 autistic adolescents and adults in Sweden to understand their attitudes towards genetic research in autism. We asked whether they think genetic research in autism is positive and beneficial, what they hope the research will achieve, and if they have any concerns. Involving the autism community in Sweden is crucial for the future of genetic research in autism, ensuring it addresses their priorities and concerns. Overall, respondents had a positive view, hoping the findings could lead to better interventions, improved quality of life, enhanced educational opportunities, and stronger support services for autistic individuals and their families. However, many were worried about potential negative consequences, such as increased discrimination and the promotion of eugenics. These findings can guide researchers in designing future studies and highlight the importance of community involvement in research.

## Introduction

Autism is a highly heterogeneous neurodevelopmental condition with childhood onset. The global prevalence is estimated to be around 1-2%, with numbers rising in recent years. More boys are diagnosed with autism compared to girls (Baxter et al., 2015; Loomes et al., 2017; Zeidan et al., 2022). Current diagnostic practices rely on behavioral assessments, caregiver interviews, and the criteria outlined in the Diagnostic and Statistical Manual of Mental Disorders (DSM-5), with no standardized biological markers yet incorporated into clinical diagnostics (American Psychiatric Association, 2013; Lyall et al., 2021; Ramirez-Celis et al., 2022).

Genetics play a dominant role in the etiology of autism. Twin studies estimate heritability between 75–97%, suggesting that while environmental factors contribute, their role is modest and unlikely to account for the rising prevalence of diagnoses (Taylor et al., 2020). Recent genomic research has revealed a complex and diverse landscape of rare and common genetic variants, showcasing the etiological heterogeneity of autism (Litman et al., 2025; Trost et al., 2022). Understanding the genetic architecture of autism aims to improve diagnostic accuracy, identify biologically meaningful subtypes, and advance precision health approaches tailored to individual needs based on genomic data.

As scientific advances in the field of autism genetics and genomics unfold, ethical and social considerations become increasingly important. Genetic research has the potential to influence how autism is perceived and experienced by the general population and, more importantly, by autistic individuals and their families (Hens et al., 2016; Manzini et al., 2021; Natri et al., 2023). In some cases, genetic studies have been paused due to concerns that the findings could fuel stigma, misunderstanding, or eugenic misuse rather than offer a tangible benefit (Sanderson, 2021). Earlier research indicates that both autistic adults and parents of autistic children generally support genetic studies when they believe the research is respectful, well-communicated, and has the potential to improve quality of life or inform support services (Ellis & Ashbury, 2023; Johannessen et al., 2016). However, some members of the autism community have expressed unease that research agendas may be misaligned with their lived experience priorities, focusing more on causation and early detection than on interventions, quality of life, or acceptance (den Houting et al., 2021; E. Pellicano et al., 2014, 2014). Studies have also highlighted a lack of diversity in who is represented in autism research, particularly with respect to gender, intellectual disability, and cultural background (Bottema-Beutel et al., 2023).

Little is known about the attitudes of the Swedish autism community toward genetic research. To address this gap, we conducted an online survey of autistic individuals and parents of autistic children to understand their perspectives. We explored their views on research into the genetic causes of autism, their willingness to participate in studies, the importance of receiving research results at both the individual and group levels, and concerns about the ethical and social implications. This study aims to center community voices in shaping future Swedish autism genetics research.

## Methods

### Survey design

We conducted two online questionnaires to gather opinions and experiences on various aspects of genetics. One questionnaire targeted parents with at least one child with an autism diagnosis, and the other questionnaire targeted adolescents (from 15 years) and adults with an autism diagnosis. The surveys were accessible online from October 12th to December 1st, 2020. The survey and data collection were done using Survey&Report version 4.3.10.5.

The overall structure and development of the survey have been described earlier (Hellquist & Tammimies, 2022). In brief, the parent survey consisted of 62 questions, while the adolescent and adult survey had 51 questions, including demographic information. Both included closed and open-ended questions, with five questions focused on genetic research used for this study (Supplementary Table 1). When developing the questionnaire, we received feedback from one parent of an autistic child, two genetic counselors, a medical doctor, and a representative from an autism organization, leading to minor revisions for clarity and brevity. Furthermore, the second round of revisions was done after piloting the questionnaire with three parents of autistic children.

### Recruitment of responders and informed consent

Recruitment for potential responders has also been described earlier (Hellquist & Tammimies, 2022) . In brief, recruitment was conducted through various online and social media channels as well as child and adolescent psychiatry clinics and habilitation centers. Informed consent was obtained after respondents read the information sheet in the online survey. The estimated time to complete the surveys was between 15 and 30 minutes. The surveys and the study were reviewed and approved by the Swedish Ethical Review Authority (dnr 2020-03291).

### Data Analysis

Data from both surveys were downloaded from the Survey&Report system and processed using Microsoft Excel, R version 4.3.2, and R Studio. The demographic information is presented using descriptive statistics for the groups separately. Demographic and survey responses differences between the parent-group and the autism-group were analyzed using the χ² test and Fisher’s exact test, depending on response size. Linear regression analyses were conducted to examine the relationship between various demographic information and concerns regarding genetic research in autism. The figure with the identified themes was made in Canva (https://www.canva.com/). A reflexive thematic analysis was conducted inductively according to Braun & Clarkés Thematic Analysis Framework when analyzing the concerns regarding genetic research in autism (Byrne, 2022). In the presentation of findings, respondents are identified by a code in parentheses at the end of each quotation (e.g., P005). In some cases, the same respondent is quoted more than once when their response illustrated multiple themes and sub-themes. The quotations are direct translations from Swedish to English, and therefore reflect participants’ own choices of terminology, ranging from identity-first language (“autistic person”) to person-first language (“person with autism”).

## Results

### Demographic information

The demographic characteristics of the 871 parent-group and the 213 autism-group have been described previously (Hellquist & Tammimies, 2022). For the current analysis, we included all respondents. We conducted subgroup analyses within the autism-group, including those who responded to the open-ended question about fears or concerns regarding genetic research (Table 1). Among parents, most respondents were women (94.6%), aged 36–45 years (48.9%), with a monthly household income of 40,000–59,999 SEK (31.7%), and university degrees (52.6%). The most commonly reported conditions among parents were exhaustion disorder (30.5%), depression (25.0%), and anxiety (24.7%).

**Table 1.** Demographic information on study respondents.

| Characteristics | Parents to children with autism (=871)<br>Number (%) | Adolescents and adults with autism who commented with a fear on genetic research<br>Number (%) |  |  |
| --- | --- | --- | --- | --- |
|  |  | Yes (n=103) | No (n=109) | All (n=212) |
| Gender |  |  |  |  |
| Woman | 824 (94.6) | 73 (70.9) | 86 (78.9) | 159 (74.7) |
| Man | 39 (4.5) | 21 (20.4) | 21(19.3) | 43 (20.2) |
| Non-binary | <5 | 6 (5.8) | <5 | 7 (3.3) |
| Other | <5 | <5 | 0 | <5 |
| Unsure | 0 | <5 | <5 | <5 |
| Prefer not to answer | <5 | <5 | 0 | <5 |
| Age |  |  |  |  |
| 15 – 18 | 0 | <5 | 7 (6.4) | 10 (4.7) |
| 19 – 25 | <5 | 12 (11.7) | 15 (13.8) | 27 (12.7) |
| 26 – 35 | 93 (10.7) | 33 (32.0) | 36 (33.0) | 70 (32.9) |
| 36 – 45 | 426 (48.9) | 25 (24.3) | 27 (24.8) | 52 (24.4) |
| 46 – 55 | 310 (35.6) | 20 (19.4) | 12 (11.0) | 32 (15.0) |
| 56 – 65 | 34 (3.9) | 8 (7.8) | 11 (10.1) | 19 (8.9) |
| >65 | <5 | <5 | <5 | <5 |
| Prefer not to answer | <5 | 0 | 0 | 0 |
| Monthly household income in SEK |  |  |  |  |
| 0 – 19 999 | 48 (5.5) | 30 (29.1) | 46 (42.2) | 77 (36.2) |
| 20 000 – 39 999 | 194 (22.3) | 28 (27.2) | 32 (29.4) | 60 (28.2) |
| 40 000 – 59 999 | 276 (31.7) | 18 (17.5) | 13 (11.9) | 31 (14.6) |
| 60 000 – 89 999 | 253 (29.1) | 16 (15.5) | 11 (10.1) | 27 (12.7) |
| > 90 000 | 83 (9.5) | <5 | <5 | 6 (2.8) |
| Prefer not to answer | 17 (2.0) | 7 (6.8) | 5 (4.6) | 12 (5.6) |
| Education |  |  |  |  |
| Compulsory school | 38 (4.4) | 10 (9.7) | 30 (27.5) | 40 (18.9) |
| Upper secondary school | 142 (16.3) | 21 (20.4) | 31 (28.4) | 52 (24.4) |
| Post upper secondary school | 83 (9.5) | 9 (8.7) | 7 (6.4) | 17 (8.0) |
| University studies | 148 (17.0) | 20 (19.4) | 13 (11.9) | 33 (15.5) |
| University degree | 458 (52.6) | 42 (40.8) | 27 (24.8) | 69 (32.4) |
| Prefer not to answer | <5 | <5 | <5 | <5 |
| Co-occurrence of other neurodevelopmental disorders |  |  |  |  |
| Autism |  |  |  |  |
| Yes | 62 (7.1) | N/A | N/A | N/A |
| No | 804 (92.3) |  |  |  |
| Prefer not to answer | 5 (0.6) | N/A | N/A | N/A |
| ADHD | 96 (11.0) | 30 (29.1) | 34 (31.2) | 64 (30.1) |
| ADD | 33 (3.8) | 21 (20.4) | 17 (15.6) | 39 (18.3) |
| Intellectual disability | <5 | <5 | <5 | <5 |
| Tourettes syndrome | <5 | <5 | <5 | 8 (3.8) |
| Dyslexia | 32 (3.7) | 9 (8.7) | 10 (9.2) | 19 (8.9) |
| Dyscalculia | 11 (1.3) | 5 (4.9) | <5 | 6 (2.8) |
| No | 710 (81.5) | 47 (45.6) | 50 (45.9) | 97 (45.5) |
| Prefer not to answer | 10 (1.2) | <5 | <5 | <5 |
| **Co-occurrence of psychiatric disorders** |  |  |  |  |
| Depression | 218 (25.0) | 62 (60.2) | 65 (59.6) | 128 (60.1) |
| Anxiety | 215 (24.7) | 58 (56.3) | 65 (59.6) | 124 (58.2) |
| Specific phobia | 17 (2.0) | 10 (9.7) | 9 (8.3) | 19 (8.9) |
| Social phobia | 48 (5.5) | 16 (15.5) | 28 (25.7) | 45 (21.1) |
| Sleeping problems | 181 (20.8) | 51 (49.5) | 47 (43.1) | 99 (46.5) |
| Exhaustion syndrome | 266 (30.5) | 34 (33) | 36 (33.0) | 71 (33.3) |
| OCD | 21 (2.4) | 13 (12.6) | 13 (11.9) | 26 (12.2) |
| Anorexia | 6 (0.7) | <5 | 8 (7.3) | 11 (5.2) |
| Bipolar disorder | 15 (1.7) | <5 | 6 (5.5) | 8 (3.8) |
| Psychosis | 0 | 0 | <5 | <5 |
| Schizophrenia | <5 | 0 | <5 | <1 |
| PTSD | 64 (7.4) | 20 (19.4) | 14 (12.8) | 34 (16.0) |
| Alcohol abuse | 6 (0.7) | 0 | <5 | <5 |
| Drug abuse | <5 | <5 | <5 | <4 |
| Other | <5 | 18 (17.5) | 5 (4.6) | 24 (11.3) |
| No | 401 (46.0) | 12 (11.7) | 16 (14.7) | 28 (13.2) |
| Prefer not to answer | <5 | <5 | <5 | <3 |

Within the autism-group, most respondents were women (74.7%), aged 26–35 years (32.9%), with a monthly income of 0–19,999 SEK (36.2%) (Table 1). Educational attainment within the autism-group included upper secondary school (24.4%) and university degrees (32.4%). %).

We also analyzed whether there were any demographic differences among the two stratified subgroups within the autism-group (Supplementary Table 2). There was a significant difference in educational attainment as those who left a comment on their concerns on genetic research in autism (n = 103) had a significantly higher proportion with university degrees (40.8%) compared to those who did not comment (24.8%, p = 0.019). The group that left a comment also included fewer individuals in the lowest income category (0–19,999 SEK: 29.1% vs. 42.2%); however, this difference was not significant (p > 0.05).

### Attitudes toward genetic research and willingness to participate

We first surveyed the respondents to determine their attitudes towards genetic research in autism. Parents were significantly more likely to report being positive towards genetic research to a very large extent (47.0% vs. 33.3%; p = 4.46e-4) (Figure 1A). Less than half of respondents in both groups believed that future research could prevent autism or lead to pharmaceutical treatments; however, the parent-group were more likely than the autism-group to expect such prevention or treatment to a large extent (30.7% vs. 22.5%; p = 2.41e-02), while the autism-group were more likely to answer not at all (25.8% vs. 11.1%; p = 5.86e-08) (Figure 1B). The majority in the autism-group expressed their concerns about potential misuse of genetic research, with 61.1% responding that they worry to a very large and a large extent (Figure 1C). The parent-group expressed more concerns in comparison to the autism-group, with a total of 67.1% expressing concern that genetic research may lead to selection against fetuses with a high likelihood of autism to a very large extent and a large extent. A significant difference emerged in concerns expressed to a large extent, with 37.9% of parents doing so compared with 26.8% of the autism-group (p = 3.09e-03, Figure 1C). In contrast, a small fraction of the autism-group expressed that they were not at all concerned, which was a significantly higher percentage than in the parent-group (9.9% and 2.3%, respectively, p = 6.16e-07). When respondents were asked whether they would be willing to participate in genetic research in autism, 70% of the respondents in the autism-group and 68.4% in the parents-group indicated a willingness to either personally participate in or allow their child to participate in research aimed at identifying the genetic causes of autism (Figure 1D).

**Figure 1.**
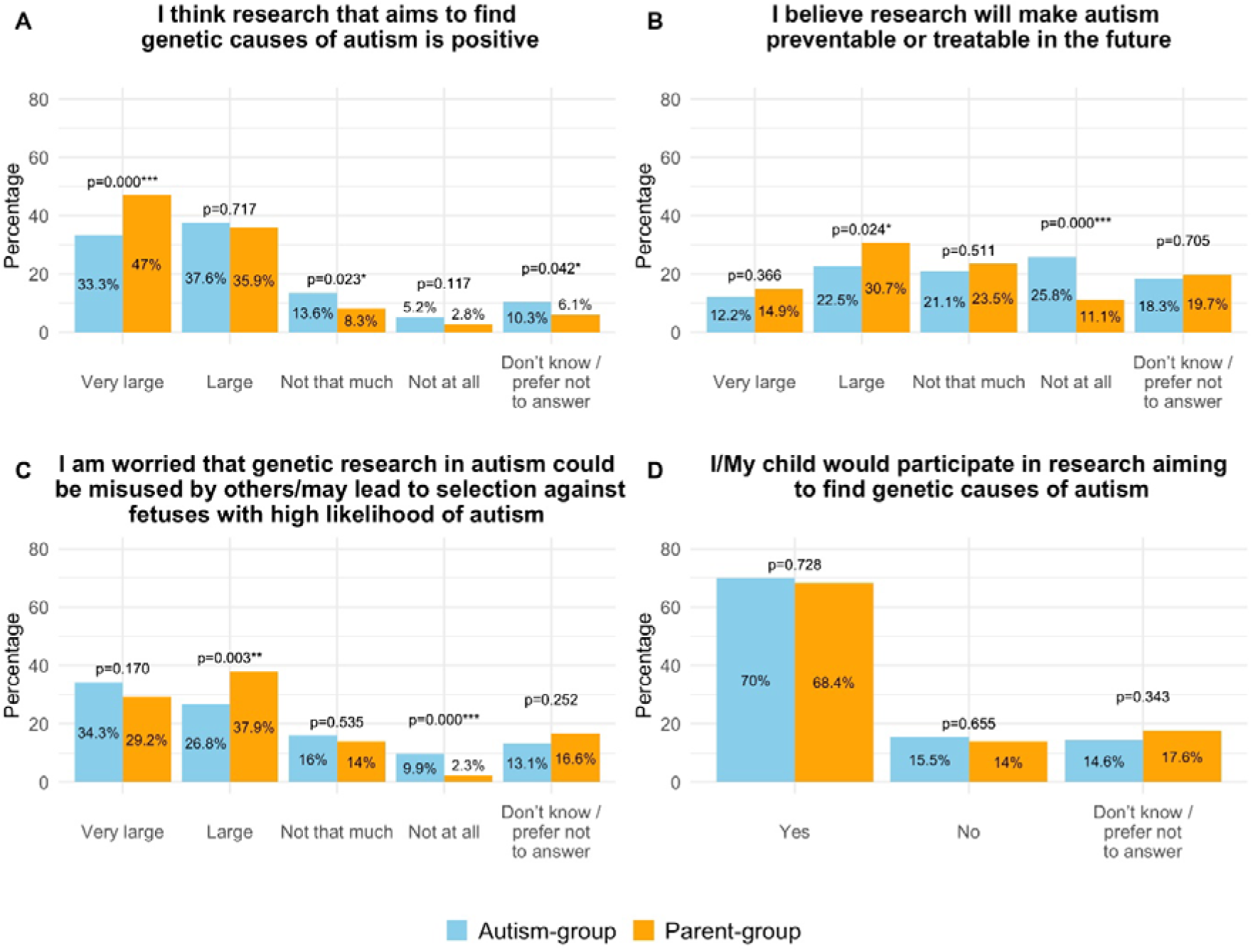
Comparison of attitudes toward genetic research in autism between autistic adolescents/young adults and parents of autistic children. Comparison of the responses between the autism-group (blue) and the parent-group (orange) across four items: (A) perceived positivity of genetic research, (B) belief that genetic research could make autism preventable or treatable, (C) concerns about potential misuse or selective practices, and (D) willingness to participate in genetic research. Items were worded slightly differently for the two groups to reflect their perspectives. Between-group differences were assessed using χ² or Fisher’s exact tests, with significant differences indicated by p-values.

### Associations between demographics and concerns about genetic research

We evaluated the potential influence of demographic factors on concerns regarding genetic research in autism using linear regression analyses. Within the autism-group, we focused on concerns about the potential misuse of genetic research in relation to income and education. We identified significant negative correlations between concerns about genetic misuse and both income (p = 0.011) and education (p = 4e-4), suggesting that respondents with lower incomes and lower levels of education expressed greater concerns. (p>0.05) (Supplementary Table 3).

For the parent-group, we analyzed whether concerns about a potential increase in pregnancy terminations due to prenatal genetic screening were associated with demographic factors. In this group, no significant associations were found with income (p>0.05) and education (p>0.05) (Supplementary Table 3).

### Respondent preferences for genetic result disclosure and research priorities

One potential benefit of participating in genetic research is access to one’s genetic information such as potential pathogenic variants. To examine this, we asked respondents about the importance of receiving individual genetic results. The majority of respondents preferred individual genetic results. As many as 85.9% in the parent-group and 90.1% of the autism-group, responded that it was important to receive individual genetic results (Figure 2A). Furthermore, a majority in both groups preferred to be informed of other secondary risks (78.6% of the parent-group and 72.9% of the autism-group) (Figure 2B). One example of such other secondary risks is the discovery of a pathogenic genetic variant linked to genetic conditions unrelated to autism, such as hereditary breast cancer.

**Figure 2.**
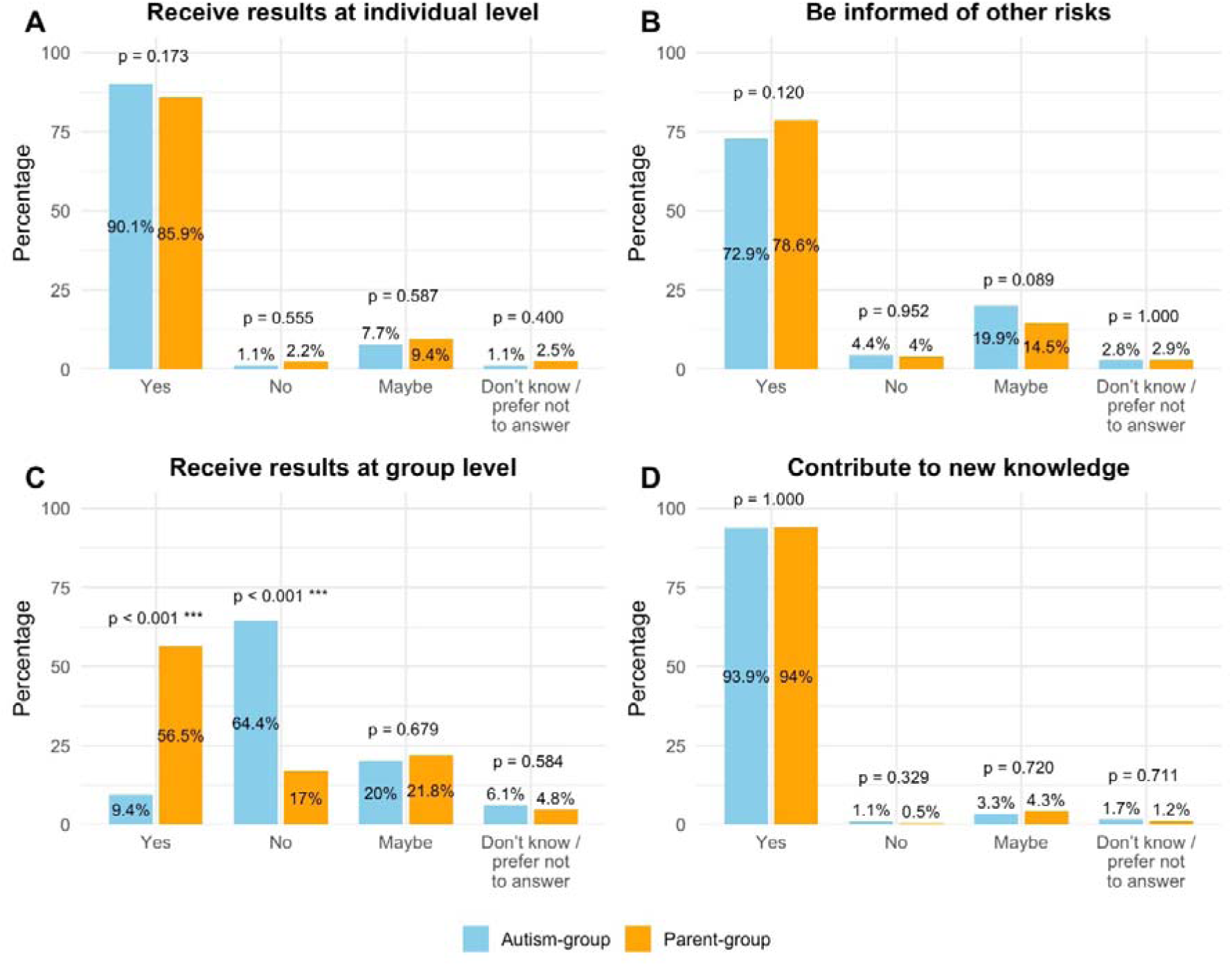
Motivation for participation in autism genetic research among autistic individuals and parents of autistic children. Comparison of the responses between the autism-group (blue) and the parent-group (orange) regarding motivations to participate in autism genetic research: (A) the return of individual-level results, (B) the return of additional genetic risks, (C) the return of only group-level results, and (D) that research would contribute to new knowledge. Between-group differences were assessed using χ² or Fisher’s exact tests, with significant differences indicated by p-values.

Preferences for group-level results differed significantly between groups. Only 9.4% of autistic respondents valued group-level findings, compared with 56.5% of parents (p = 1.72e-29). Conversely, 64.4% of autistic adults stated they would not participate if only group-level results were provided, whereas only 17% of parents reported the same (p = 3.02e-38) (Figure 2C).

More than 90% of the autism-group and parent-group showed motivation to participate in genetic research that could generate knowledge potentially benefiting others with autism, including interventions to improve daily life, enhance quality of life, and provide better educational and family support (Figure 2D).

### Thematic Analysis of Concerns Regarding Genetic Research in Autism

As previously described, most autism-group respondents expressed substantial apprehension (Figure 1C). Nearly half (n=103; Supplementary Table 2) provided comments, which underwent qualitative thematic analysis, resulting in four overarching themes (Figure 3)

**Figure 3.**
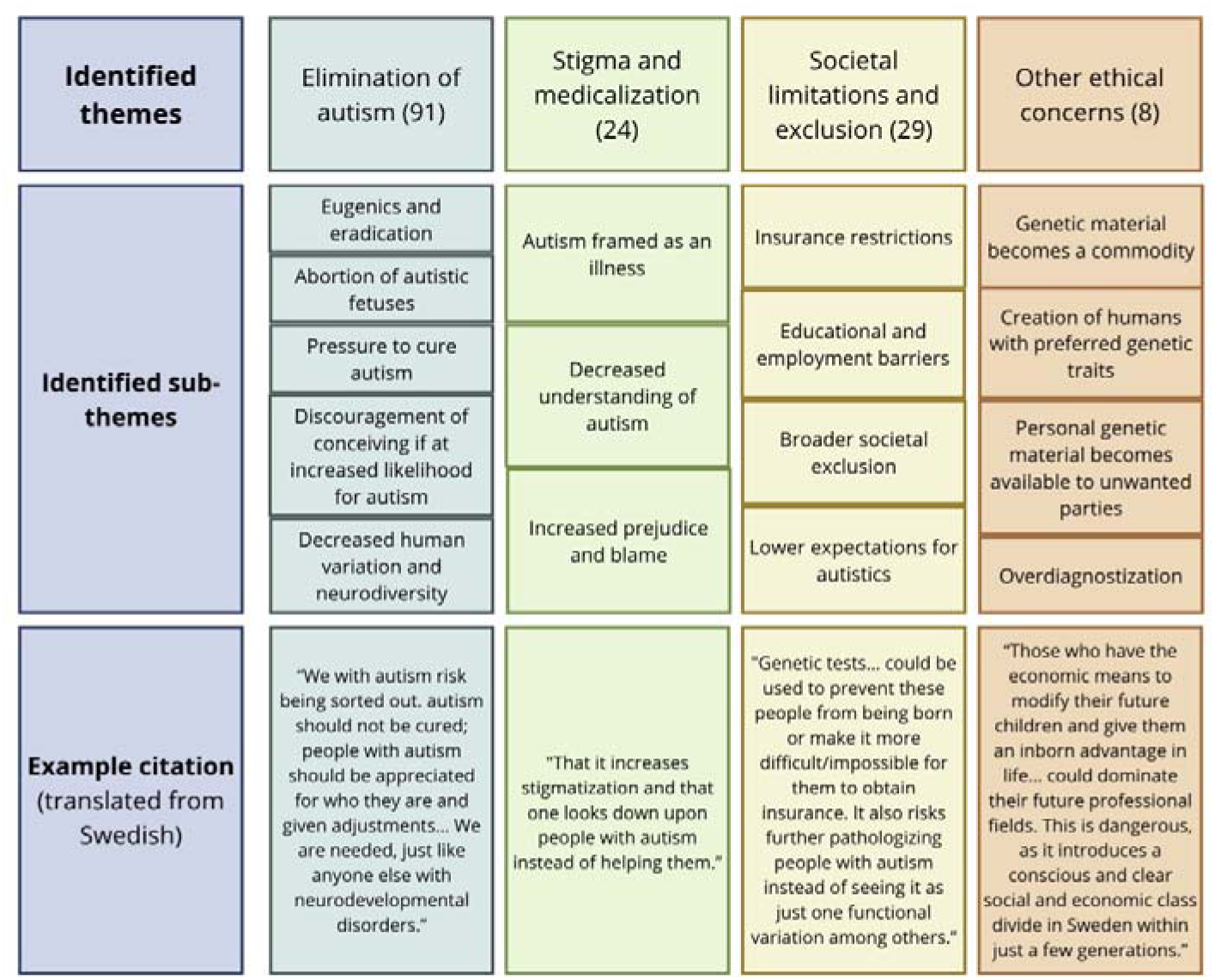
Identified concerns regarding potential negative implications of autism genetic research in the autism-group from thematic analysis of the open-ended question on the topic. Numbers in parentheses indicate the frequency of theme occurrence across the dataset.

#### Theme 1: Elimination of autism

The most frequent concern was the potential for genetic research to lead to the elimination of autism. This included concerns about eugenics and eradication of autistics, abortion of autistic fetuses and increased pressure to cure autism.

#### Sub-theme 1.1: Eugenics and eradication

Many respondents feared that genetic research could revive eugenic practices, framing autism as a defect rather than a valued human variation. Historical parallels were common in the comments, with some comparing prenatal screening to Nazi racial hygiene: “*When people start checking fetal genes to eliminate all deviations, it is honestly eugenics… If this discussion existed about any other group of people, everyone would call it Nazi ideas and say no thanks*” (P025). Another respondent reflected, “*I would not want to change who I am… I hope genetic research finds new ways to help us, not erase us*” (P056).

This fear extended to eradication concerns, with some warning of catastrophic misuse under oppressive regimes:

“*I am afraid we will all be sent to gas chambers to be eradicated in a society that wants more efficiency and fewer people who are not seen as useful. I am afraid it could happen sooner than we think*” (P039).

#### Sub-theme 1.2: Abortion of autistic fetuses

A prominent concern among respondents was that genetic research could be used to justify the abortion of autistic fetuses, erasing autistic lives before birth. Several envisioned a future of “genetic planning,” where deviations from neurotypical are eliminated:

“*Genetic planning, that fetuses showing deviations from what NT [neurotypical] people consider normal will be aborted. Kallocain warning…”* (P010).

Another expressed this even more starkly: “*Abort Aspies [autistics], see living Aspies as walking failed abortions who should have been killed*” (P094).

These concerns were closely tied to diversity and societal loss:

“*We with Asperger’s together with high IQ are overrepresented among for example, Nobel Prize winners and scientists, and therefore we should not be aborted away*” (P103).

#### Sub-theme 1.3: Pressure to cure autism

Many respondents strongly rejected the notion of medically ‘curing’ autism, emphasizing it as a core part of identity rather than an illness. As one explained, , “*I do not want medication or attempts to cure everyone with autism… There is nothing wrong with having autism, and stigma increases if it is seen as a curable disease, when it is not even a disease - just a different way of functioning*” (P087). Or as another respondent put it, “*Autism should not be cured; people with autism should be appreciated for who they are and given adjustments… We are needed, just like anyone else with neurodevelopmental disorders*” (P005).

Others viewed attempts to eliminate autism through research or family planning as profoundly dehumanizing:

“*I think it is a completely horrible idea to try, through genetic research and possible family planning, to eliminate or cure autism. I WANT to keep my autism. I suspect at least one of my children has autism, and I wouldn’t want it in any other way. What I wish is less stigma and greater understanding* (P074)”. Several warned that promoting a cure risk legitimizing efforts to erase autistic people rather than support them.

#### Sub-theme 1.4: Discouragement of conceiving if at increased likelihood for autism

For some, the implications of genetic knowledge extended beyond abortion or cure, raising concerns that potential parents could be discouraged, or even prevented, from having children. Concerns about discrimination and echoes of historical abuses were explicitly mentioned: “*Discrimination, racial thinking in a transferred sense, forced sterilization*” (P099). This sense of coercion was echoed in other responses: “*That future or new parents refrain from having children, or that a parent is blamed or not allowed to become a parent*” (P067). The possibility of genetic data being used politically or coercively was a recurring concern, as highlighted in the extended reflection from P062, who warned that registers of genetic risk could be used to “*weed out carriers of these genes*,” that parents might feel “*defective if society thinks they should not have children when they know about the genetic risk*,” and that such practices could ultimately reinforce stigma and inequality.

#### Sub-theme 1.5: Decreased human variation and neurodiversity

Finally, respondents expressed concern that the elimination of autistic individuals through genetic testing and abortion would diminish human diversity. As one respondent put it, “*If this can already be mapped genetically during the fetal stage, many fetuses will be aborted, which leads to a lack of diversity where autistic people are actively selected away*” (P101).

Others saw this not just as a loss for autistic people, but for society as a whole: “*To select us away could lead to a society with a lower degree of neurodiversity and therefore a less resilient and open society*” (P071).

#### Theme 2: Stigma and medicalization

Respondents also expressed concern that genetic research could increase stigma by framing autism as an illness rather than a natural variation, potentially shifting public perception toward viewing it as a treatable illness.

#### Sub-theme 2.1: Autism framed as an illness

Many respondents expressed concerns that genetic testing could reinforce the view of autism as a medical defect. One explained that genetic tests giving definitive answers about autism would risk “*further disease-stamping people with autism instead of seeing it as a functional variation among others*” (P083).

Others echoed this concern, worrying that autism might be more often defined as an illness needing treatment. One noted that if autism “*starts to be counted as a disease instead of a functional ‘impairment*,’” then people with less obvious symptoms such as women or those with autism level 1 might find it harder to receive a diagnosis, thereby losing access to support and adaptations. The same respondent pointed out that healthcare could increasingly frame autism as something to be “*cured with medication,” implying that autism itself was “something bad*,” though they added that for individuals with very severe forms of autism, medical research and treatments could indeed be beneficial (P068).

#### Sub-theme 2.2: Decreased understanding of autism

Respondents also worried that advances in genetics would not improve understanding but would instead deepen misconceptions. One respondent feared that genetic results “*in the wrong hands*” would not increase understanding. The respondent stressed that “*there are already far too many prejudices, assumptions, and fear from people who think that we on the autism spectrum are odd*,” although autistic people often bring important strengths (P002)”.

Some respondents were concerned that autism would increasingly be seen as something negative or shameful. One remarked that genetic testing could “*increase stigmatization and make people look down on those who have autism instead of helping*” (P066). Another commented that autism is often “*seen as a bad thing, that you are a worse person*,” but they emphasized that it also carries strengths that should not be overlooked (P060).

#### Sub-theme 2.3: Increased prejudice and blame

A related concern was that prejudice would grow, particularly if genetic testing encouraged society to treat autistic lives as less valuable. One respondent drew a direct comparison to prenatal testing for Down syndrome, explaining that while the respondent had been offered the CUB test *[combined ultrasound scan and blood test]* in pregnancy, they declined. The respondent reflected that as many parents choose abortion when expecting a child with Down syndrome, it “*sends signals that DS [Down syndrome] children are unwanted*.” The respondent worried that if autism were similarly tested for prenatally, stigma could grow.

Finally, some respondents were concerned that stigma could extend to parents themselves, who might be blamed for having autistic children if prenatal testing enabled autism to be “screened out.” One expressed the risk that autism would increasingly be seen as “wrong,” leading to “blaming parents” or pushing them toward abortion “as with Down syndrome,” alongside attempts to “cure” autism altogether (P096).

Another respondent warned that support and resources could be reduced if parents were held responsible for choosing to continue pregnancies. They imagined parents being told, “*Blame yourselves for having a child with a diagnosis,*” and feared that the autistic community would become “*a more vulnerable and much smaller group in society*” (P079).

#### Theme 3: Societal limitations and exclusion

Respondents also worried that genetic information could lead to exclusion across insurance, education, healthcare, and employment, not only for autistic individuals but also for those with elevated genetic vulnerability to autism, resulting in reduced opportunities and inadequate societal support.

#### Sub-theme 3.1: Insurance restrictions

A recurring concern was that access to insurance could be severely restricted. As one respondent put it, “*Obtaining insurance [would be more difficult, resulting in] an even more challenging life; poor economy and poor social life*” (P047). Others envisioned a scenario where genetic information would lead to exclusion altogether: “*Genetic tests… could be used to prevent these people from being born or make it more difficult/impossible for them to obtain insurance. It also risks further pathologizing people with autism instead of seeing it as just one functional variation among others*” (P083). Another expressed similar worries: “*I am afraid it will lead to a large number of abortions of fetuses with a high genetic probability for autism, and that insurance companies could start excluding people with certain genetic variants*” (P081).

#### Sub-theme 3.2: Educational and employment barriers

Respondents feared that genetic labeling could limit access to schools, training, or jobs: “*Worse insurance, worse education, not being accepted into certain programs, being excluded from certain tasks or professions, abortions*” (P041). For some, the diagnosis of autism had already meant restrictions on employment. One explained, “*After more than 20 years as a daily driver without any incidents, now that I have a diagnosis, every other year I must present a certificate to the Swedish Transport Agency to prove that I can still drive… Now suddenly I live with a label in many contexts and am limited in my choice of profession - jobs I could handle perfectly well before, I am now suddenly not allowed to apply for*” (P021).

#### Sub-theme 3.3: Broader societal exclusion

Respondents also raised serious concerns about healthcare and broader social participation. Some were concerned that registers of genetic vulnerabilities could be misused for political purposes and introduce broader forms of societal exclusion. One worried that “*data ends up in the wrong hands… with the rise of SD [Swedish Democrats] this feels extra relevant, claiming that someone does not need support because they don’t have genetic autism, or conversely assuming someone must be a certain way from the beginning… increased inequality in health and medical care… leading to mental illness and premature death*” (P062). Others reflected on how existing systemic biases could be amplified, with one writing, “*In a capitalist unregulated society, individuals can easily be harmed by how information is used. Knowledge is good if it helps, but bad if it is used to discriminate. I am especially worried that opportunities will be denied to people on the basis of a diagnosis… If I lived in the US, I would be worried that healthcare, insurance, etc. would become more expensive, while wages and job opportunities decreased*” (P030).

#### Sub-theme 3.4: Lower expectations for autistics

One respondent voiced the concern that genetic testing could lead to reduced expectations being placed on autistic children. They worried that society might respond by “*choosing more abortions, choosing away their children, and having low expectations of their children*” (P061). While the respondent did not explicitly link this to prenatal testing, the implication was that if autism were identifiable already in the fetal stage, it might be framed as a limitation rather than a potential.

#### Theme 4: Other ethical concerns

A minority of respondents raised additional ethical concerns, including the commodification of genetic material, the creation of humans with preferred traits, the misuse of personal genetic data, and overdiagnosis. Some feared a future where wealthy families could purchase genetic advantages, creating social and economic divides: *those who have the economic means to modify their future children and give them an inborn advantage in life… could dominate their future professional fields. This is dangerous, as it introduces a conscious and clear social and economic class divide in Sweden within just a few generations*” (P037), or where future society might “genetically cultivate superhumans and sort out defective fetuses” (P052). Others worried about genetic information falling into the wrong hands, as one noted “*leak out into cyberspace so anyone can access it*” (P104). One respondent highlighted risks of overdiagnosis and its putative consequnces, “*a diagnosis = limitations*” and should only be applied “*when you need extra support or treatment, not automatically to everyone meeting the criteria*” (P021).

#### Recognizing strengths and the need for societal support

Finally, although it was not part of the survey questions, some respondents felt it was important to highlight the positive qualities of autistic individuals and the need for societal support. Respondents emphasized strengths and abilities, as one explained, “*Autism is not something that should be eradicated! There are many positive sides, such as logical thinking, focus, accuracy, loyalty, and a strong sense of justice, which are important assets to society. However, greater understanding is needed in society regarding how to address and manage the difficulties that may come with it*” (P009). In total, sixteen respondents highlighted positive qualities of autistic individuals, while nine respondents emphasized the importance of societal support and understanding.

## Discussion

The field of genetic research in autism has been expanding, yet there remains a notable scarcity of studies exploring how individuals within the autistic community perceive such (Fletcher-Watson, Larsen, et al., 2019; Kapp et al., 2013; E. Pellicano et al., 2014). Our study contributes to the understanding of these attitudes within a Swedish setting. Overall, our findings reveal a cautiously optimistic attitude toward genetic research within the autistic community. Both autistic individuals and parents recognized the potential benefits of such research, including improved diagnostic accuracy, better understanding of co-occurring conditions, and the possibility of interventions that alleviate daily challenges. These views are consistent with prior studies in the UK and other countries (Asbury et al., 2024; Fletcher-Watson, Larsen, et al., 2019; Nicolaidis, 2012; E. Pellicano et al., 2014).

Respondents emphasized that research should enhance autistic well-being rather than aim to eliminate or prevent autism, aligning with studies showing community priorities centered on health, support, and inclusion (Roche et al., 2021; Warner et al., 2019). While some saw potential value in genetic testing and counseling (Byres et al., 2023; Ellis & Ashbury, 2023), many warned that such information could reinforce ableist narratives or eugenic misuse.

A major concern was potential misuse of genetic research. Approximately 85% of respondents were concerned that research could lead to prenatal screening and selective terminations. Similar apprehensions have been reported elsewhere, even contributing to the suspension of UK genetic study and sparking autistic-led protests around the world (Frazier et al., 2018; Pugsley et al., 2025; Sanderson, 2021). Similar views have also been raised for clinical genetic testing by the autistic community (Hendry et al., 2025). Comparisons were frequently drawn to prenatal screening for Down syndrome, which has reduced the number of births (de Graaf et al., 2021). Respondents also worried about discrimination based on genetic predispositions, consistent with evidence of pervasive exclusion in employment and education (Botha & Frost, 2020; Cleary et al., 2023; Gonzales, 2022; Praslova, 2021; Sasson et al., 2017).

We found that individuals with lower socioeconomic status or education were more likely to express concerns about the negative societal consequences of genetic research. This may reflect perceived vulnerability or mistrust regarding how genetic information might be used. These findings highlight the ethical responsibility of researchers to anticipate and address social implications (Mezinska et al., 2021; Natri et al., 2023).

A recurring theme in our study was the ambivalence expressed by respondents. While they acknowledged the potential benefits of genetic research, many remained skeptical about its capacity to meaningfully improve lives without addressing systemic societal barriers. For instance, the prevalence of ableism in healthcare and research has long shaped the autism research agenda toward prevention rather than inclusion and support (Botha & Cage, 2022; Miller & Levine, 2013). This tension underscores the need for participatory research models that center autistic voices and prioritize their lived experiences (Fletcher-Watson, Adams, et al., 2019; Hobson et al., 2023; L. Pellicano et al., 2013).

As genetic research in autism progresses, it needs to be a priority that the research agenda reflects the priorities and values of the autistic community. Historically, the field has been criticized for perpetuating ableist assumptions that autism is a “deficit” requiring prevention or eradication (Botha & Cage, 2022; Cervantes et al., 2021; Gewin, 2023). Many researchers and advocates now promote a strengths-based, neurodiversity-affirming framework that emphasizes inclusion, well-being, and societal contributions of autistic individuals. The neurodiversity movement views autism as a natural variation rather than a disorder to be cured, challenging traditional medical models and highlighting the importance of acceptance and accommodation (Kapp et al., 2013; Robertson, 2010). This perspective is also reflected among parents of nonverbal and minimally verbal autistic children, who express interest in basic scientific and genomic research but emphasize that such work should aim to improve understanding and support for autistic individuals and not to eradicate or cure autism (Asbury et al., 2024).

The emerging reality of embryo selection and genetic enhancement amplifies the ethical issues raised by respondents with fears for the commodification of genetics, the creation of “preferred” traits, and the misuse of genetic information. These concerns align closely with current debates on polygenic embryo screening and germline editing (Fox et al., 2024; Lencz et al., 2022; Visscher et al., 2025). These technologies, while marketed for health benefits, risk normalizing selective reproduction and widening social inequalities, and creating a future where genetic privilege determines opportunity. These concerns are not hypothetical but increasingly relevant as reproductive genetics moves toward consumer-driven enhancement.

### Limitations

This study offers the first study of attitudes toward genetic research in autism within the Swedish autistic community, but several limitations should be acknowledged. The data is self-reported, which introduces risks of recall inaccuracies, misunderstanding of survey items, and biases linked to who chose to respond. Non-response bias is also possible, as participants may differ from non-participants.

The sample consisted predominantly of women in both the parent- and autism-groups, which limits the generalizability of the findings and may capture primarily female perspectives.

Some respondents, particularly within the autism group, interpreted that some elements of the survey implied that autism is undesirable or should be eliminated, which they associated with eugenic implications. These interpretations, reflected in seventeen qualitative responses, may have influenced how some participants answered the survey. Greater involvement of autistic community members in the development of questions may have reduced these misunderstandings. Incomplete responses were also common, potentially due to the survey’s length or the perceived irrelevance of certain questions.

An additional limitation is the inconsistency between the survey given to parent-group and the autism-group. For example, only the parent-group survey addressed concerns about pregnancy termination, while the autism-group survey asked more broadly about misuse of genetic results. The parent-group also lacked a comment box for elaboration. These differences hinder direct comparison and limit parents’ ability to express more nuanced views; some parents also wished for additional open-ended and multiple-choice questions on their views regarding research on potential pharmacological treatments for autism.

Finally, recruitment primarily occurred through online channels, which could have biased the sample toward individuals more engaged with online information or toward parents of children with greater needs. This recruitment method may also have contributed to a lower response rate among autistic adolescents and adults than among parents, suggesting that our recruitment strategy may not have fully captured this group’s perspectives.

## Conclusion

In conclusion, while the autistic community in Sweden appears broadly supportive of genetic research, they explicitly express the need to have access to their data. Despite being positive, there are persistent concerns regarding potential negative consequences. Transparent and ethical engagement with these concerns is essential to ensure responsible advancement within the field. Involving the autistic community at every stage of the research process, from study design, data interpretation, and dissemination, will ensure that their priorities and apprehensions are fully incorporated (Fletcher-Watson, Adams, et al., 2019; Fletcher-Watson, Larsen, et al., 2019; Hobson et al., 2023; Natri et al., 2023). Such participatory approaches not only strengthen the ethical foundation of research but also amplify the likelihood that findings will meaningfully serve the needs of autistic individuals, improve their quality of life, and reduce discrimination. Ultimately, research done *with* the community, rather than *on* the community, holds the greatest potential for lasting positive impact.

## Ethical consideration

The study and questionnaires were reviewed by the Swedish Ethical Review Authority (dnr.2020-03291). The respondents gave an informed consent for the study and for the use of their anonymous answers.

## Data Availability

The data will be accessible, after necessary clearances, through the Swedish National Data Service's (SND) research data catalogue.

## Acknowledgements

The authors thank all the parents and autistic adolescents, and adults for their time and valuable information. In addition, the authors thank organizations and individuals who helped us to spread the information about the questionnaire, including Attention, Autism- och Aspergerförbundet in Stockholm, Hjärnfonden, Prima Psykiatri, Ung Autism, and Riksförbundet Sällsynta Diagnoser. The authors also thank Dr MaiBritt Giacobini and Dr Lynnea Myers for valuable comments and help with the study.

## CRediT roles

Samuelle Fajutrao Falk: Writing – original draft, Formal analysis

Anna Hellquist: Conceptualization, Data curation, Investigation, Methodology, Resources, Supervision, Validation, Visualization, Writing – review & editing

Kristiina Tammimies: Conceptualization, Data curation, Funding acquisition, Investigation, Methodology, Resources, Software, Supervision, Validation, Visualization, Writing – review & editing

