## Supplementary Tables for "A survey-based study investigating opinions on genetic research among Swedish autistic individuals and parents of autistic children"

Fajutrao Falk et al.

### Supplementary Table 1. Survey questions on opinions on genetic research included in this study

<table>
<tr><td colspan="4">Survey for autistic adults</td></tr>
<tr><td></td><td colspan="2">Question</td><td>Response options</td></tr>
<tr><td>1</td><td colspan="2">I think research that aims to find genetic causes of ASD is positive.</td><td>To a very large extent<br>To a large extent<br>Not that much<br>Not at all<br>Don’t know / prefer not to answer</td></tr>
<tr><td>2</td><td colspan="2">I would participate in research aiming to find genetic causes of ASD.</td><td>Yes<br>No<br>Don’t know / prefer not to answer</td></tr>
<tr><td rowspan="5">3</td><td colspan="2">If I participate in genetic research on ASD, I would like to:</td><td rowspan="5">Yes<br>No<br>Maybe<br>Don’t know / prefer not to answer</td></tr>
<tr><td>A.</td><td>Receive the results at the individual level (that is, the results of the analysis conducted specifically for you).</td></tr>
<tr><td>B.</td><td>Be informed if researchers find that I have an increased risk for any other disease or condition besides ASD.</td></tr>
<tr><td>C.</td><td>Only receive results at the group level (that is, not the results of the analysis conducted specifically for you).</td></tr>
<tr><td>D.</td><td>Contribute to new knowledge that may hopefully help other children, adolescents, and adults with ASD in the future.</td></tr>
<tr><td>4</td><td colspan="2">I believe that research on ASD will lead to ASD being preventable in the future, or that symptoms of ASD can be treated through new medications or other interventions.</td><td>To a very large extent<br>To a large extent<br>Not that much<br>Not at all<br>Don’t know / prefer not to answer</td></tr>
<tr><td>5</td><td colspan="2">I am worried that the results from research identifying genetic variants that increase the likelihood of ASD could be misused by others.<br><br>If yes, please describe what you think would be a misuse in the field below.</td><td>To a very large extent<br>To a large extent<br>Not that much<br>Not at all<br>Don’t know / prefer not to answer</td></tr>
<tr><td colspan="4">Survey for parents to autistics</td></tr>
</table>

| | | | |
|---|---|---|---|
| 1 | I think research that aims to find genetic causes of ASD is positive. | | To a very large extent<br>To a large extent<br>Not that much<br>Not at all<br>Don’t know / prefer not to answer |
| 2 | I would allow my child to participate in research aiming to find genetic causes of ASD. | | Yes<br>No<br>Don’t know / prefer not to answer |
| 3 | If my child were to participate in genetic research on ASD, it is important that: | | Yes<br>No<br>Maybe<br>Don’t know / prefer not to answer |
| | A. | We receive the results at the individual level (that is, the results of the analysis conducted specifically for your child). | |
| | B. | We are informed if it is found that the child has an increased risk for another disease or condition besides ASD. | |
| | C. | We receive the results only at the group level (that is, not the results of the analysis conducted specifically for your child). | |
| | D. | The research leads to new knowledge that may hopefully help other children with ASD in the future. | |
| 4 | I believe that current genetic research (mapping of risk genes/variants for ASD) may lead to a future where fetuses showing a high likelihood of developing ASD could be selected against through prenatal diagnostics. | | To a very large extent<br>To a large extent<br>Not that much<br>Not at all<br>Don’t know / prefer not to answer |
| 5 | I believe that research on ASD will lead to ASD being preventable in the future, or that symptoms of ASD can be treated through new medications or other interventions. | | To a very large extent<br>To a large extent<br>Not that much<br>Not at all<br>Don’t know / prefer not to answer |

**Supplementary Table 2.** Demographic differences between respondents who left a comment and those who did not in the autism-group.

| **Characteristics** | Comment with a fear regarding genetic research | | P-value* |
|---|---|---|---|
| | Yes (n=103) Number (%) | No (n=109) Number (%) | |
| **Gender** | | | |
| Woman | 73 (70.9) | 86 (78.9) | 0.23 |
| Man | 21 (20.4) | 21 (19.3) | 0.97 |
| Non-binary | 6 (5.8) | <5 | 0.06 |
| Unsure | <5 | <5 | 1 |
| Prefer not to say | <5 | 0 | 0.98 |
| **Age** | | | |
| 15 – 18 | <5 | 7 (6.4) | 0.43 |
| 19 – 25 | 12 (11.7) | 15 (13.8) | 0.92 |
| 26 – 35 | 33 (32.0) | 36 (33.0) | 1 |
| 36 – 45 | 25 (24.3) | 27 (24.8) | 1 |
| 46 – 55 | 20 (19.4) | 12 (11.0) | 0.09 |
| 56 – 65 | 8 (7.8) | 11 (10.1) | 0.82 |
| > 65 | <5 | <5 | 0.23 |
| **Income** | | | |
| 0 – 19 999 | 30 (29.1) | 46 (42.2) | 0.07 |
| 20 000 – 39 999 | 28 (27.2) | 32 (29.4) | 0.84 |
| 40 000 – 59 999 | 18 (17.5) | 13 (11.9) | 0.34 |
| 60 000 – 89 999 | 16 (15.5) | 11 (10.1) | 0.33 |
| > 90 000 | <5 | <5 | 0.63 |
| Prefer not to answer | 7 (6.8) | 5 (4.6) | 0.69 |
| **Education** | | | |
| Primary school | 10 (9.7) | 30 (27.5) | 0.0017 |
| Secondary school | 21 (20.4) | 31 (28.4) | 0.23 |
| Post-secondary school | 9 (8.7) | 7 (6.4) | 0.71 |
| University studies | 20 (19.4) | 13 (11.9) | 0.19 |
| University degree | 42 (40.8) | 27 (24.8) | 0.019 |
| Prefer not to say | <5 | <5 | 1 |
| **Comorbidity with other neurodevelopmental disorders** | | | |
| ADHD | 30 (29.1) | 34 (31.2) | 0.72 |
| ADD | 21 (20.4) | 17 (15.6) | 0.56 |
| Intellectual disability | <5 | <5 | 0.63 |
| Tourettes syndrome | <5 | <5 | 1 |
| Dyslexia | 9 (8.7) | 10 (9.2) | 1 |
| Dyscalculia | 5 (4.9) | <5 | 0.21 |

| | | | |
|---|---|---|---|
| No | 47 (45.6) | 50 (45.9) | 0.88 |
| Prefer not to answer | <5 | <5 | 1 |
| **Comorbidity with other psychiatric disorders** | | | |
| Depression | 62 (60.2) | 65 (59.6) | 0.95 |
| Anxiety | 58 (56.3) | 65 (59.6) | 0.88 |
| Specific phobia | 10 (9.7) | 9 (8.3) | 0.87 |
| Social phobia | 16 (15.5) | 28 (25.7) | 0.14 |
| Sleeping problems | 51 (49.5) | 47 (43.1) | 0.47 |
| Exhaustion syndrome | 34 (33) | 36 (33.0) | 1 |
| OCD | 13 (12.6) | 13 (11.9) | 1 |
| Anorexia | <5 | 8 (7.3) | 0.27 |
| Bipolar disorder | <5 | 6 (5.5) | 0.34 |
| Psychosis | 0 | <5 | 0.51 |
| Schizophrenia | 0 | <5 | 1 |
| PTSD | 20 (19.4) | 14 (12.8) | 0.27 |
| Alcohol abuse | 0 | <5 | 0.15 |
| Drug abuse | <5 | <5 | 0.67 |
| Other | 18 (17.5) | 5 (4.6) | **0.0063** |
| No | 12 (11.7) | 16 (14.7) | 0.70 |
| Prefer not to answer | <5 | <5 | 1 |

*

### Supplementary Table 3. Association between education and income with concerns regarding genetic research in autism.

| Variable | Estimate | Std. Error | t-value | p-value |
|---|---|---|---|---|
| Autism-group | | | | |
| Income | -0.170 | 0.0656 | -2.59 | **0.011** |
| Education | -0.177 | 0.0491 | -3.60 | **0.0004** |
| Parent-group | | | | |
| Income | 0.0171 | 0.0281 | 0.610 | 0.54 |
| Education | -0.0230 | 0.0232 | -0.991 | 0.67 |
